# Validation of the Hindi Version of the Bronchiectasis Health Questionnaire in a Hindi-speaking Indian population

**DOI:** 10.64898/2026.02.04.26345330

**Authors:** Prem Thilak Palani, Shrawan Raut, Prayas Sethi, Rahul Krishnan Gopalakrishnan, Ved Prakash Meena, Sanjeev Sinha, Naveet Wig, Animesh Ray

## Abstract

**Background:** Bronchiectasis is a debilitating respiratory condition characterized by chronic cough with expectoration of thick sputum. It accounts for significant morbidity and mortality, especially when associated with exacerbations. Assessing the health-related quality of life (HR-QoL) of patients with bronchiectasis is important to ascertain the impact of the disease on day-to-day life, as well as to gauge the effect of targeted interventions. Conventionally used methods for assessing HR-QoL such as the St. George’s Respiratory Questionnaire (SGRQ) are time-consuming and have limitations in day-to-day application. The Bronchiectasis Health Questionnaire (BHQ) is a novel, compact tool used for assessing the HR-QoL, and has been validated for use in Korean and Turkish populations.

**Methods:** We attempted to develop and validate the Hindi version of the Bronchiectasis Health Questionnaire (BHQ) in Indian adults with bronchiectasis. We assessed the correlation between the Hindi BHQ (H-BHQ) scores and other measures of lung health including the Hindi version of the COPD Assessment Tool (H-CAT), pulmonary function tests and the bronchiectasis severity index (BSI). In addition, we assessed the correlation between the H-BHQ scores and the number of exacerbations and hospital admissions in the previous year.

**Results:** A total of 145 subjects with bronchiectasis were included. The mean (± SD) H-BHQ total score was 49.10 ± 10.3. The H-BHQ score correlated well with the H-CAT score (Correlation coefficient -0.6534, p value < 0.0001) and the mMRC scale (Correlation coefficient of -0.4459,p value < 0.0001). The H-BHQ score also had a moderate correlation with the number of exacerbations and low correlation with hospital admissions in the previous year, with correlation coefficients of -0.4193 (p < 0.0001) and -0.3030 (p < 0.0001), respectively. The correlation between the H-BHQ and the Bronchiectasis Severity Index (BSI) score was weak (Correlation coefficient of -0.3012, p value < 0.01).

**Conclusion:** The H-BHQ offers a simple and convenient method to assess the HR-QoL in patients with bronchiectasis, and correlates well with other measures of respiratory health, including the H-CAT, the mMRC score and the number of exacerbations and hospital admissions in the previous year.

## Introduction

Bronchiectasis is a clinical condition characterized by chronic cough with expectoration of thick sputum. Pathologically, it features abnormal dilation and thickening of the bronchi due to a known or unknown insult. It affects a wide range of ages from the very young to the elderly. While in children it is often the result of a congenital or genetic condition (such as cystic fibrosis or primary ciliary dyskinesia – PCD), the pathogenesis in adults usually involves chronic airway injury secondary to tobacco smoke exposure or previous infections including but not limited to tuberculosis. Bronchiectasis is an important cause of mortality; US and British observational studies suggest 5-year mortality rates of around 12% ^1, 2^. From Asia, in a study of approximately 217000 individuals^3^ comprising subjects with CT-diagnosed bronchiectasis, subjects with bronchiectasis had a higher all-cause mortality (1608.8 per 100,000 person-years,95% CI: 1531.5–1690.0) than controls (133.5 per 100,000 person-years; 95% CI, 124.1–143.8; *P*⍰< ⍰0.001). The bronchiectasis group had higher respiratory (aHR, 3.49; 95% CI, 2.21–5.51) and lung cancer-related (aHR, 3.48; 95% CI, 2.33–5.22) mortality risks than subjects without bronchiectasis. In addition to contributing to mortality, bronchiectasisexerts a significant toll on quality of life and healthcare expenditure costs; for example, healthcare expenditure on bronchiectasis in the United States has been studied to be up to 14.8 billion USD per year.

Assessment of the health-related quality of life (HRQOL), also known as the health status, has been an important component of the management of subjects with bronchiectasis primarily to assess the impact of the disease and to guide treatment targets. A large number of questionnaires have been used to assess HRQOL in bronchiectasis, and nine of these were analysed in a 2015 systematic review^4^. Although these questionnaires were found to have good psychometric properties but there was only a weak to moderate association between HRQOL and objective outcome measures such as FEV1% predicted, exercise capacity and extent of bronchiectasis on chest imaging. The St. George’s Respiratory Questionnaire (SGRQ) is the most commonly used tool to asses HRQOL in trials involving diseases of the lung; it is self-administered and contains 50 items and 76 weighted responses divided into three components: Symptoms, Activity, and Impacts. In a study of 111 subjects with bronchiectasis, the SGRQ had decent correlation with objective analysis of lung function^5^. However, while appropriate for research, the extensive nature of the SGRQ is time-consuming (on average, it requires about 578 seconds for assessment^6^), limiting its use in day-to-day patient care. The Leicester Cough Questionnaire (LCQ) is another widely used questionnaire assessing quality of life in subjects with chronic respiratory conditions, but it has only been validated for use in subjects with unexplained chronic cough, but not in bronchiectasis subject where other symptoms like dyspnea or fatigue are commonly present^7^.

The Bronchiectasis Health Questionnaire (BHQ) was developed specifically for use in subjects with bronchiectasis; it contains 10 items and generates an overall score. The internal consistency of the BHQ and its convergent validity with the SGRQ and lung function were confirmed in a study of 206 subjects^8^. In addition, BHQ scores also correlated well with the number of exacerbations and hospital admissions in the previous year. However, since it is a self-administered questionnaire, the BHQ needs translation into local languages which needs to be studied for internal consistency, validity and reliability before it can be used as a tool in public healthcare. The BHQ questionnaire, first validated in 2017, has been translated in more than 11 languages and has been extensively in many studies on bronchiectasis. However, till now, the questionnaire has not been validated in a bronchiectasis cohort (of heterogenous aetiology) among a large Hindi-speaking population, the fourth-commonest language spoken globally^9^. This study was designed to validate the Hindi version of BHQ in Hindi speaking subjects suffering from bronchiectasis of diverse causes.

## Materials and Methods

### 1. Study subjects

We included subjects aged 18 years or older who had clinic-radiological features compatible with bronchiectasis and without any history of present or past exacerbation within the last one month^10^. Pregnant and lactating females were excluded. Baseline demographic and clinical details (including previous pulmonary infections, significant comorbidities, severity of symptoms and history of exacerbations) were collected; results of pulmonary function testing (PFT) at baseline were also performed. The subjects were systematically evaluated for the cause and severity of bronchiectasis. Besides eliciting history of past infections including pulmonary tuberculosis (microbiologically proven/clinic-radiologically diagnosed), the subjects were also evaluated for their immunoglobulin levels, IgE and IgG *Aspergillus*, PICADAR (PrImary CiliARy DyskinesiA Rule) score^11^, and sweat chloride test if there was a childhood history suggestive of cystic fibrosis. Next the severity of disease was assessed by Bronchiectasis Severity Index^12^. The data was entered by a trained personnel who screened the relevant data with the help of the authors (PT or AR). The radiological severity was assessed by a trained thoracic radiologist with more than 15 years of experience in assessing thoracic CT scans.Ethical clearance was taken from Institute Ethics Committee and informed consent was taken from every subject before enrolment (IECPG-399/20.07.2023).

### 2. Methods

#### 1) BHQ Translation

The original BHQ was translated into Hindi by the help of a team of language experts and subject experts and cross-verification by the senior author (AR).The appropriateness of the translated version was confirmed by presenting the translated version to subjects who were able to comprehend the questions and able to answer them. The Hindi version is given in the supplementary material.

#### 2) Validation

The Hindi questionnaire was self-administered by bronchiectasis subjects. The H-BHQ was validated by investigating the relationship between the H BHQ and the H-CAT scores. We also investigated the relationship between the H-BHQ scores and other variables of the mMRC dyspnea scale, lung function, exacerbations and the bronchiectasis severity index (BSI) score. The degree of correlation was graded according to conventional cut-offs; 0-0.1: Negligible correlation, 0.1 -0.38: weak correlation, 0.4-0.69: Moderate correlation, 0.7-0.89: Strong correlation, 0.9-1.0: very strong correlation^13^.

#### 3) Sample size estimation

Assuming an item to response ratio of 1:10, at least 100 subjects of bronchiectasis was planned to be included in this validation study^14,15^.

#### 4) Statistical analysis

Correlation was assessed using the Pearson’s correlation coefficient (r) for parametric variables and Spearman’s correlation coefficient (⍰) for non-parametric variables. All data are presented as mean and standard deviations (or median and inter-quartile ranges if non-parametric) for continuous variables and as frequencies and percentages for qualitative variables. A p value of < 0.05 was considered statistically significant.

## 3. Results

### 1. Subjects

A total of 145 subjects with bronchiectasis were included. The mean (± SD) age was 38.91 years, and 52.4% of the subjects were female. 79.3% of the subjects were never smokers, and 55.17% of the subjects had a history of pulmonary tuberculosis. The median forced expiratory volume in 1 second (FEV1) percent and forced vital capacity (FVC) (percentage predicted) were 53.12 ± 22.01 and 61.14 ± 19.73 respectively. The mean FEV1/FVC percent predicted was 86.45 (± 16.61). More than half of the subjects did not have an exacerbation of bronchiectasis in the previous year. The rest of the subject characteristics are summarized in the table below:

|  |  |
| --- | --- |
| Age (years) | 38.91 ± 12.97 |
| Male (%) | 69 (47.6%) |
| BMI (kg/m<sup>2</sup>) | 21.07 ± 4.62 |
| **Spirometry** |  |
| FEV1 (L) – 123 subjects | 1.16 (0.84-1.77) |
| FEV1 (%) – 123 subjects | 53.12 ± 22.01 |
| FVC (L) – 123 subjects | 1.86 ± 0.73 |
| FVC (% predicted) | 61.14 ± 19.73 |
| FEV1/FVC (% predicted) | 86.45 ± 16.61 |
| **Comorbidities** |  |
| Hypertension | 12 (8.28%) |
| Diabetes Mellitus | 7 (4.83%) |
| Asthma | 52 (35.86%) |
| Chronic Kidney Disease | 1 (0.69%) |
| **Smoking history** |  |
| Never smokers | 115 (79.3%) |
| Ever smokers | 26 (17.93%) |
| Current | 4 (2.76%) |
| **Alcohol** |  |
| Never | 122 (84.14%) |
| Ex | 16 (11.03%) |
| Current | 7 (4.83%) |
| **Previous tubercular lung disease** | 79 (54.48%) |
| mMRC |  |
| 0 | 33 (22.76%) |
| 1 | 17 (11.72%) |
| 2 | 64 (44.14%) |
| 3 | 27 (18.62%) |
| 4 | 4 (2.76%) |
| Exacerbations in previous year |  |
| 0 | 76 (52.41 %) |
| 1 | 25 (17.24 %) |
| 3 | 21 (14.48 %) |
| 4 | 6 (4.14 %) |
| 5 | 8 (5.52 %) |
| 6 | 9 (6.21%) |
| Hospital admissions in previous year |  |
| 0- 122 | 122 (84.14 %) |
| 1- 9 | 9 (6.21 %) |
| 2- 4 | 4 (2.76 %) |
| 3- 2 | 2 (1.38 %) |
| 4- 2 | 2 (1.38 %) |
| 5- 6 | 6 (4.14 %) |
| **Sputum bacterial culture positivity during the last one year (109)** | **35 (32.11 %)** |
| *Pseudomonas aeruginosa* | 15 |
| *Klebsiella pneumoniae* | 5 |
| *Escherichia coli* | 1 |
| *Streptococcus pneumoniae* | 4 |
| *Haemophilus influenzae* | 10 |
| *Staphylococcus aureus* | 1 |
| *Acinetobacter baumannii* | 1 |
| *Aspergillus*-specific IgE (kUA/L) (124 subjects) | 0.04 (0.01-0.23) |
| *Aspergillus*-specific IgG (mGA/L) (121 subjects) | 15.1 (7.05 – 35.8) |
| IgA (mg/dL) (109 subjects) | 354 (281-438) |
| IgG (mg/dL) (109 subjects) | 1652 (1382-1894) |
| IgM (mg/dL) (108 subjects) | 135.5 (91.5-194.5) |
| **Hindi BHQ (H-BHQ) score** | **49.10 ± 10.30** |
| **Hindi CAT (H-CAT) score** | **16.22 ± 7.89** |
| **Bronchiectasis Severity Index (BSI) Score (117 subjects)** | **4 (2-6)** |

### 2. Correlation between H-BHQ and other clinical measurements

The mean (± SD) H-BHQ total score was 49.10 ± 10.3. The H-BHQ score showed a moderate correlation with the H-CAT score, with a Pearson correlation coefficient of - 0.6534 (p < 0.0001). The correlation between the H-BHQ score and the mMRC dyspnea scale was assessed using a Spearman rank correlation coefficient (since mMRC is an ordinal variable); H-BHQ correlated moderately with the mMRC scale with a correlation coefficient of -0.4459 (p < 0.0001). The H-BHQ also had a moderate correlation with the number of exacerbations and a weak correlation with hospital admissions in the previous year, with correlation coefficients of -0.4193 (p < 0.0001) and -0.3030 (p < 0.0001), respectively. In addition, we also assessed the correlation between the HBQ score and the Bronchiectasis Severity Index (BSI) score; which showed a weak correlation with a coefficient of -0.3012 (p < 0.01).

Of 109 subjects who provided sputum samples, there was no statistically significant difference in the H-BHQ scores between subjects whose specimen did not grow any organism on culture (mean H-BHQ 49.04 ± 9.82) and those which yielded growth (mean H-BHQ 47.11 ± 11.51) (p value = 0.33). There were no differences in H-BHQ scores between subjects found to be colonized with *Pseudomonas aeruginosa* and those who were not.

On assessment of spirometry parameters, the H-BHQ score had a weak positive correlation with the FEV1/FVC (percent predicted) (r = 0.1317), but this was not statistically significant (p value 0.1466). The score also had a weak positive correlation with the FVC (percent predicted) (r= value 0.1990), and this correlation was statistically significant (p value 0.03).

**Fig. 1:**
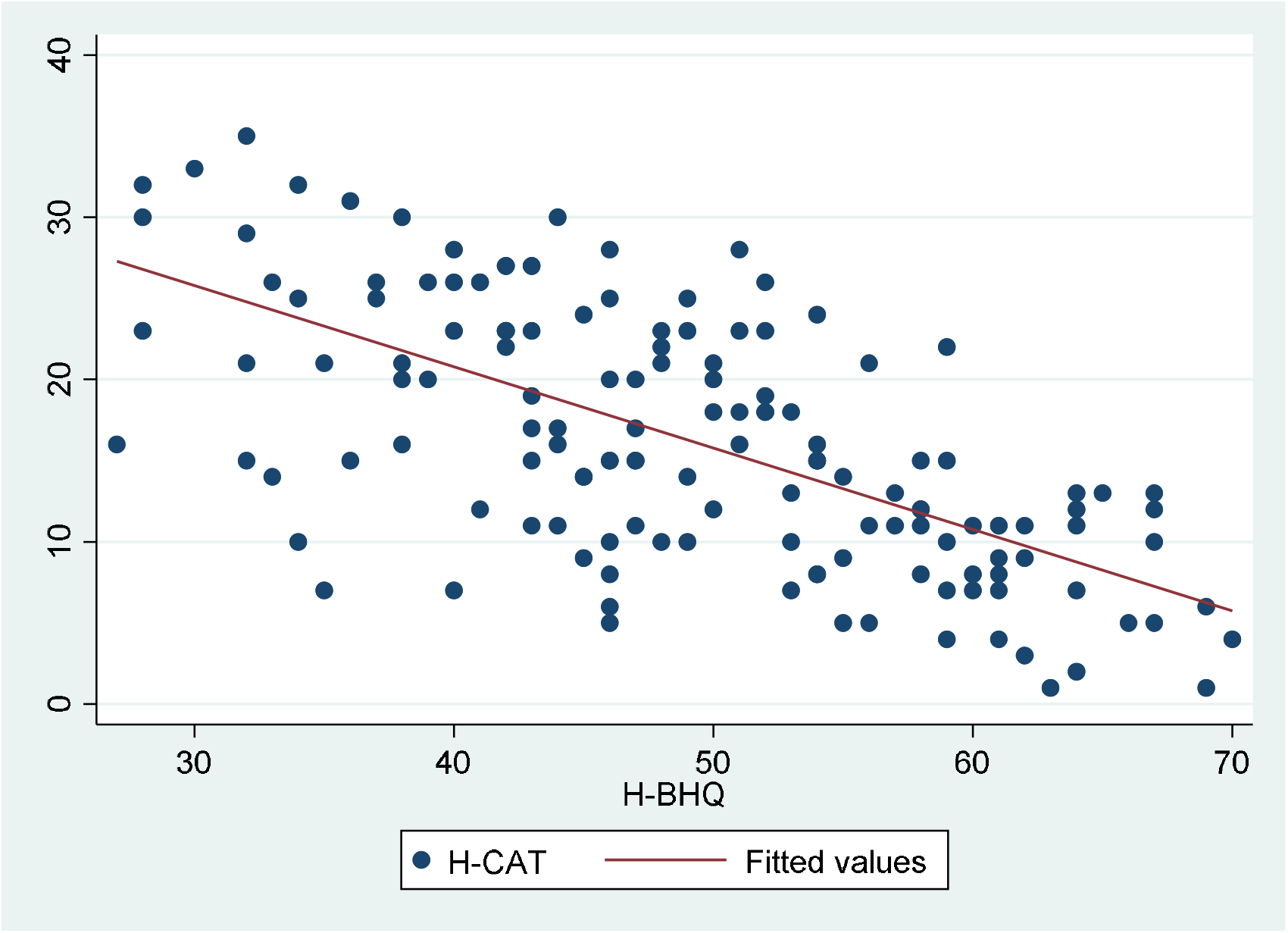
Scatter plot showing correlation between H-BHQ and H-CAT scores

**Fig. 2:**
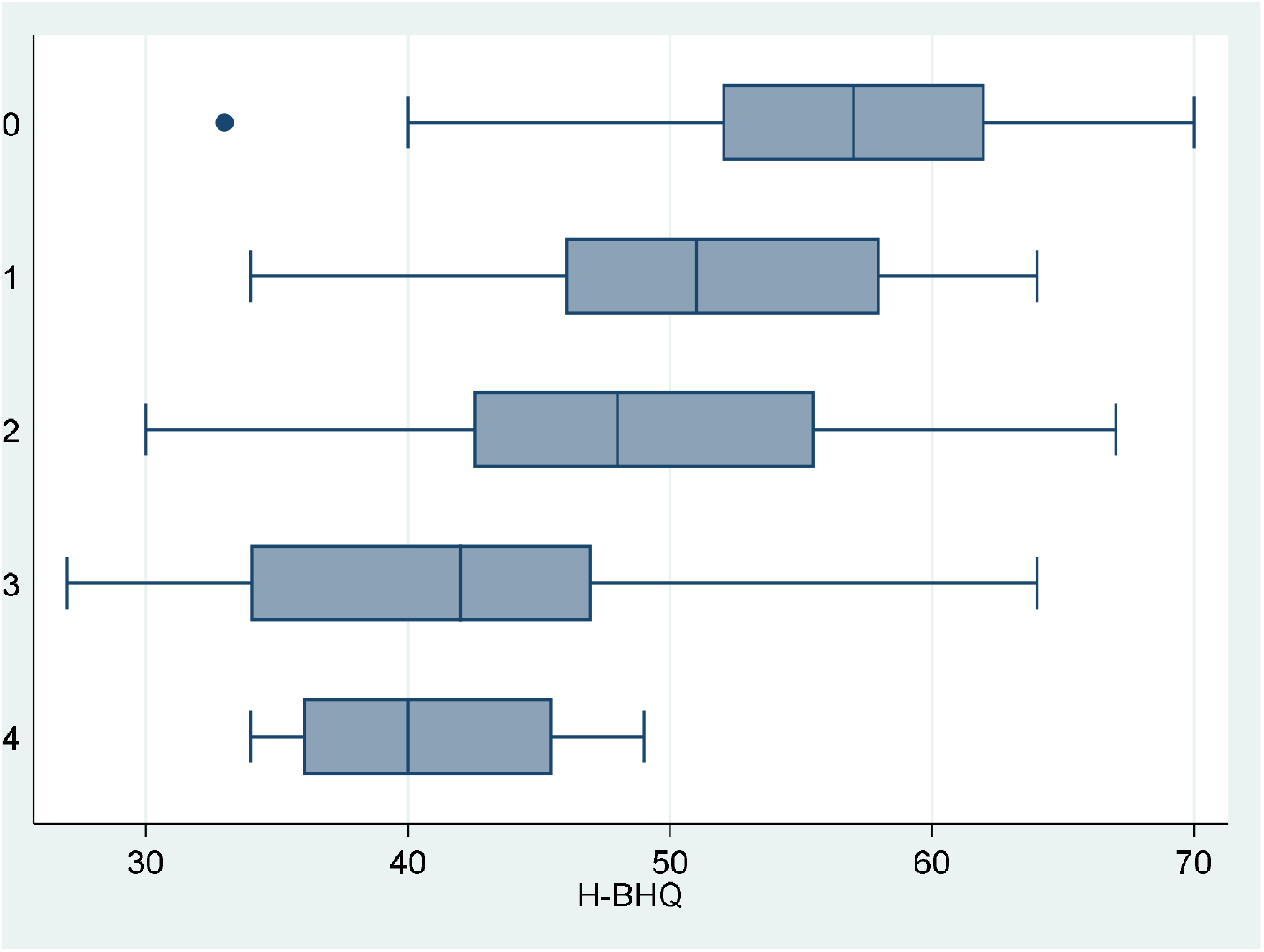
Boxplot showing distribution of HBQ scores across the 5 mMRC score groups

**Fig. 3:**
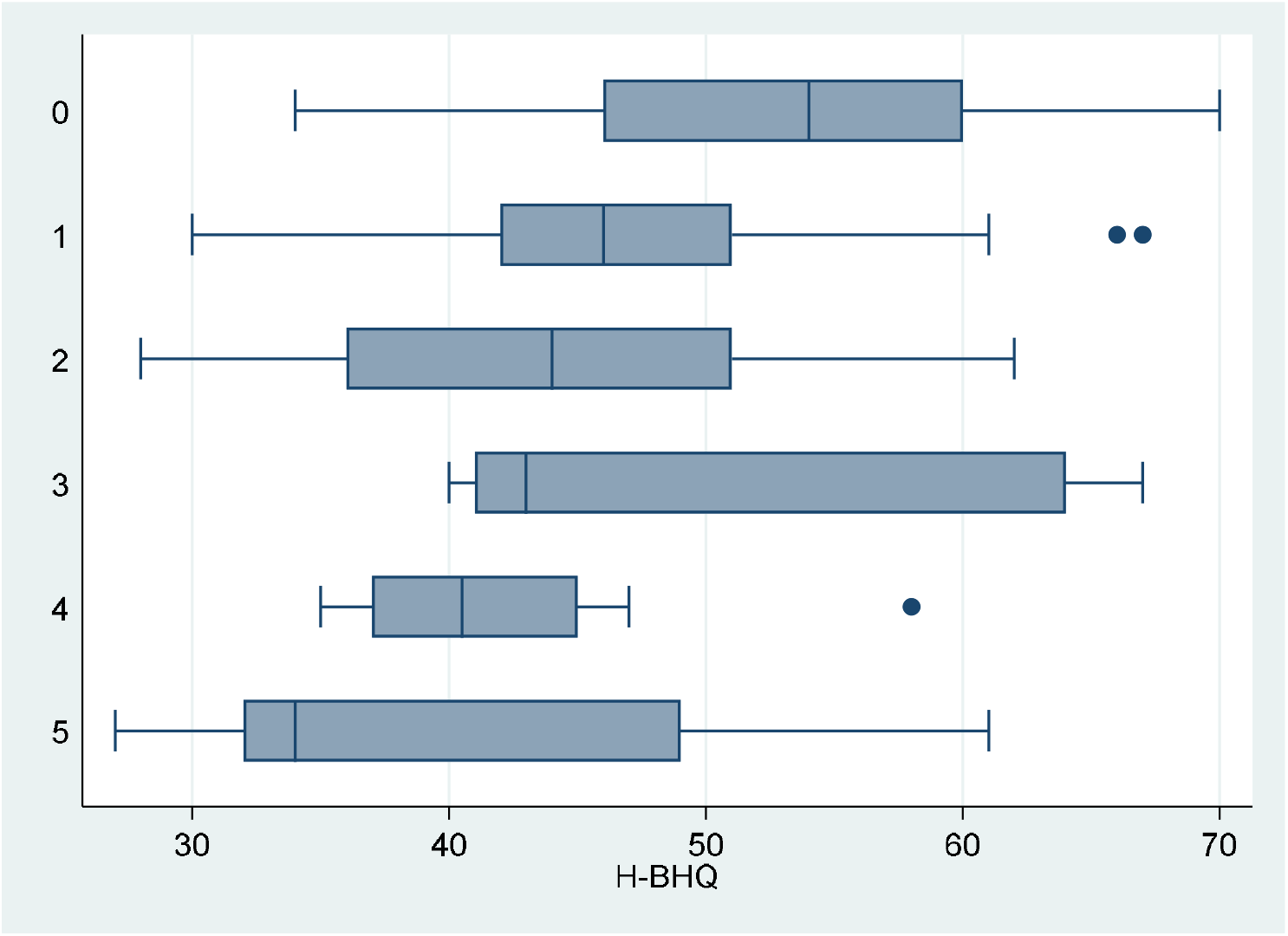
Boxplot showing distribution of HBQ scores according to the number of exacerbations in the previous year

**Fig. 4:**
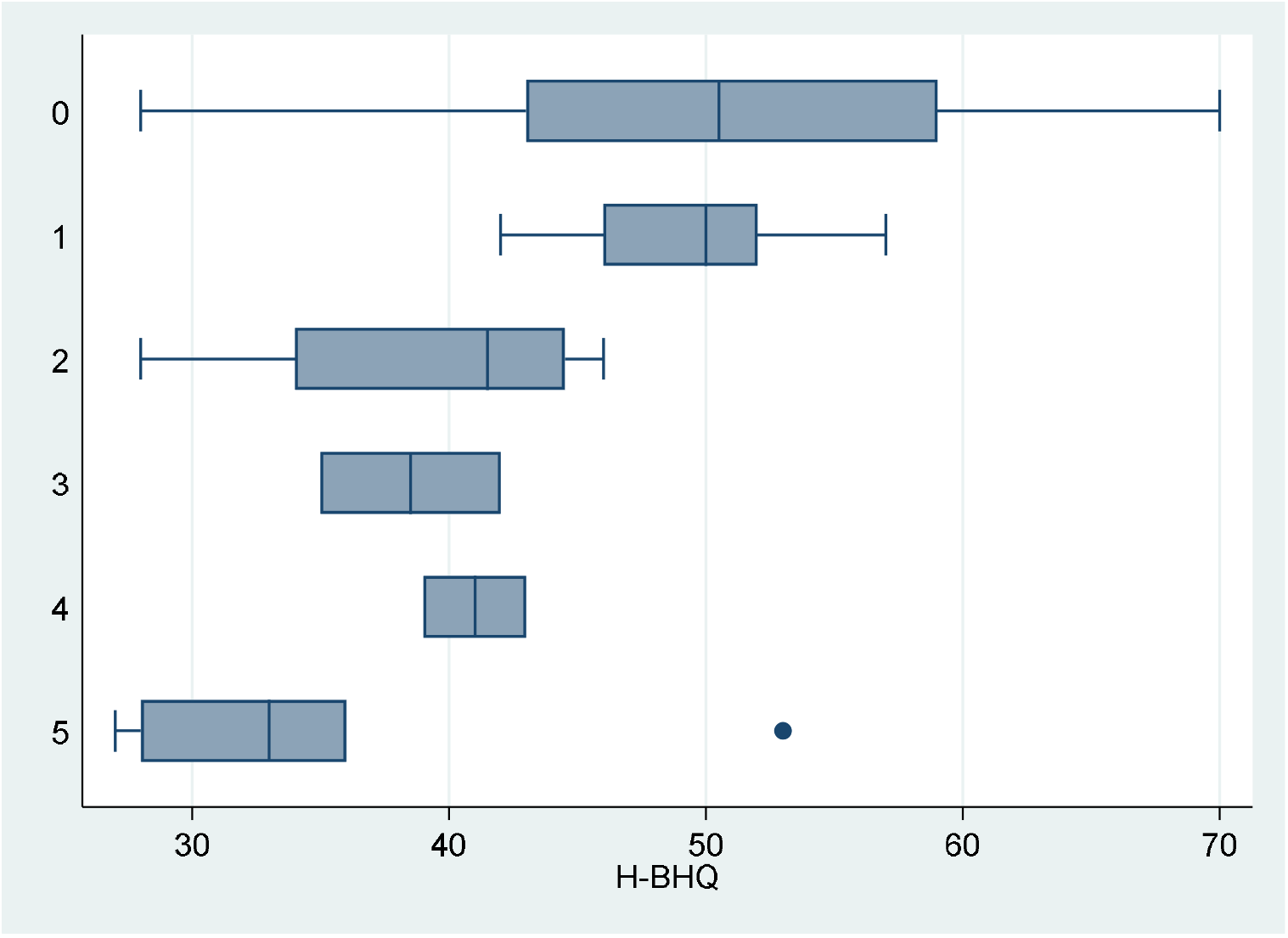
Boxplot showing distribution of HBQ scores according to the number of admissions in the previous year

**Fig. 5:**
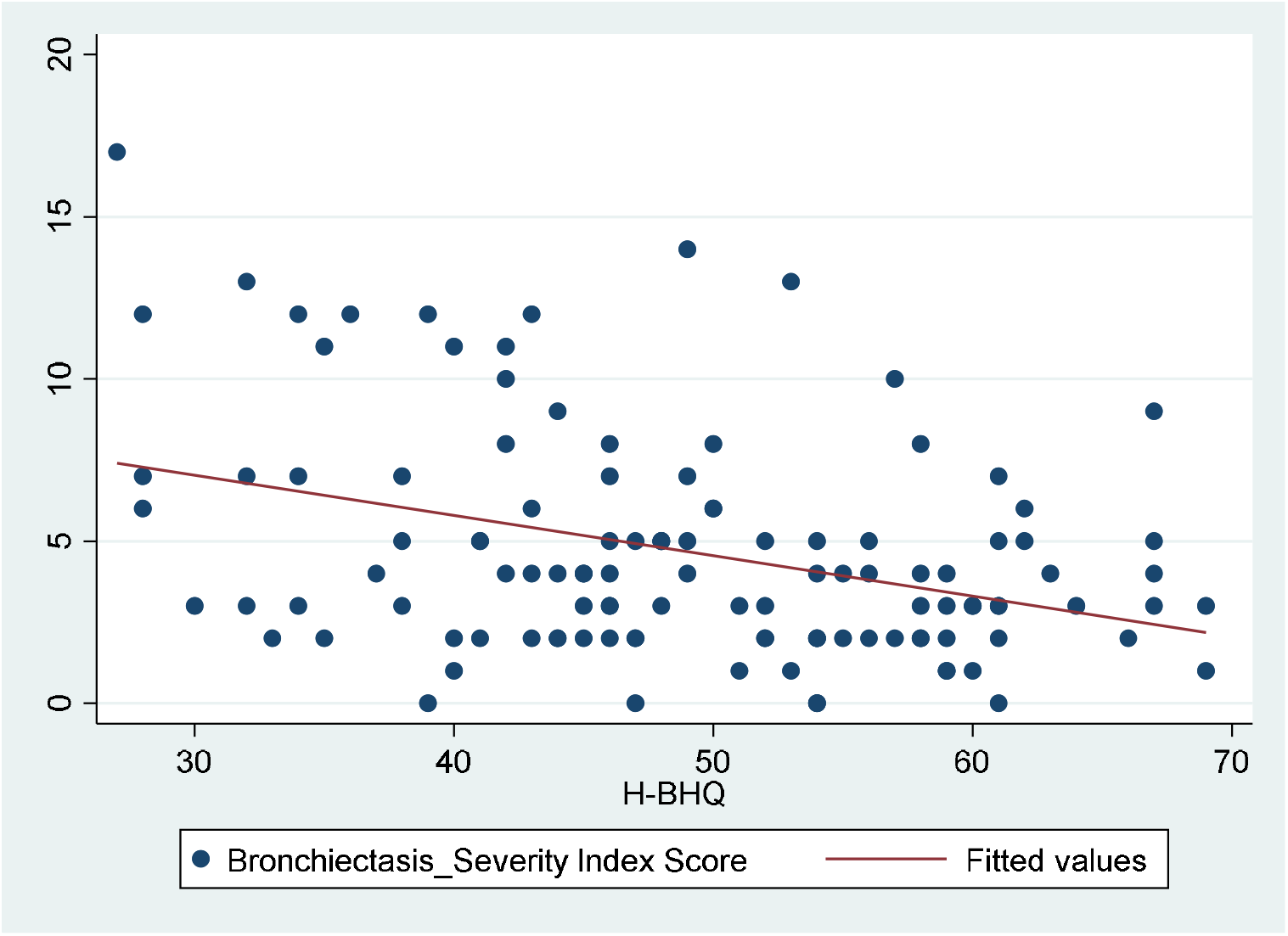
Scatter plot showing the relationship between the HBQ scores and the BSI scores

**Fig. 6:**
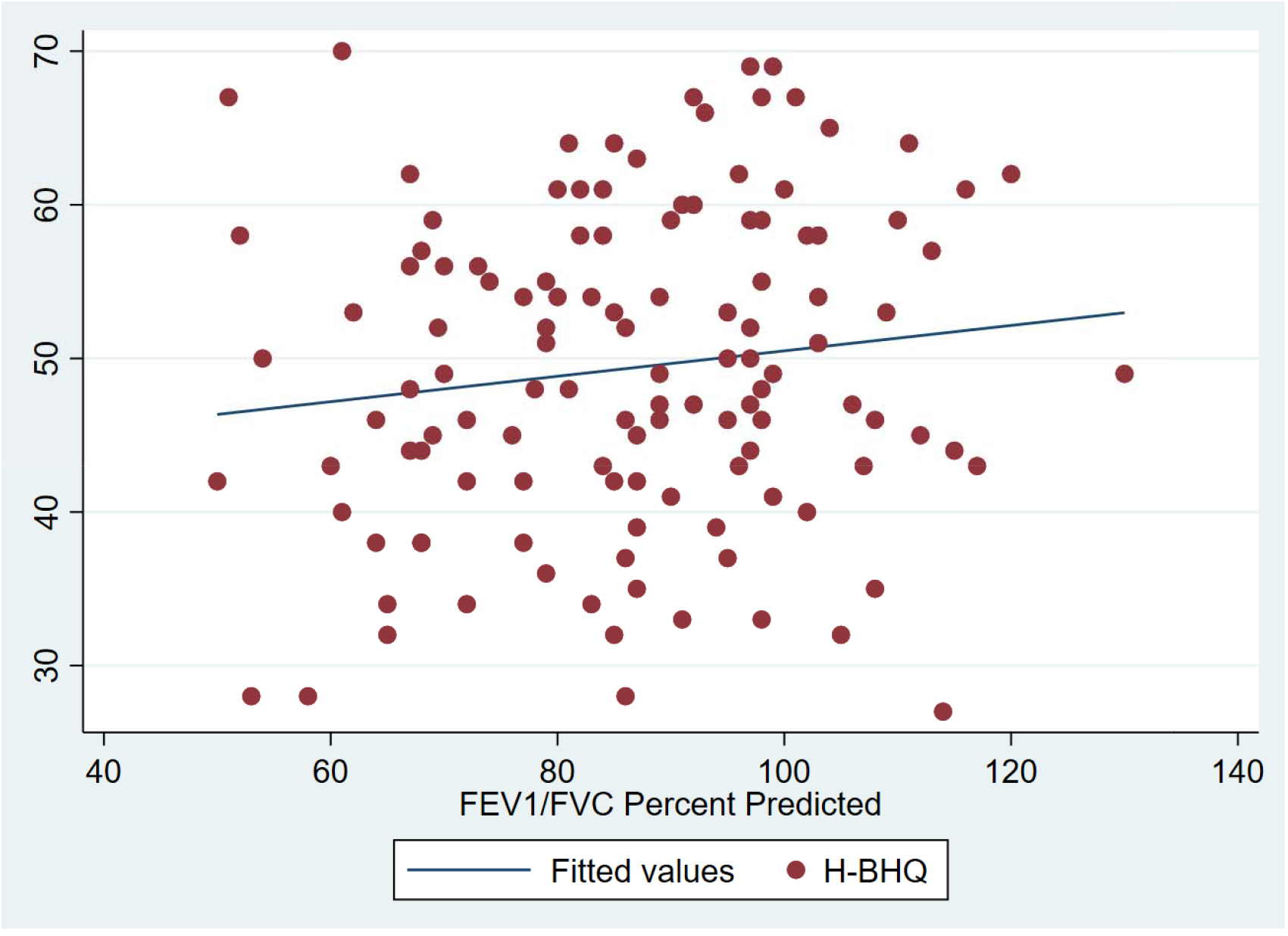
Scatter plot showing the relationship between H-BHQ score and the FEV1/FVC (percent predicted) values

**Fig. 7:**
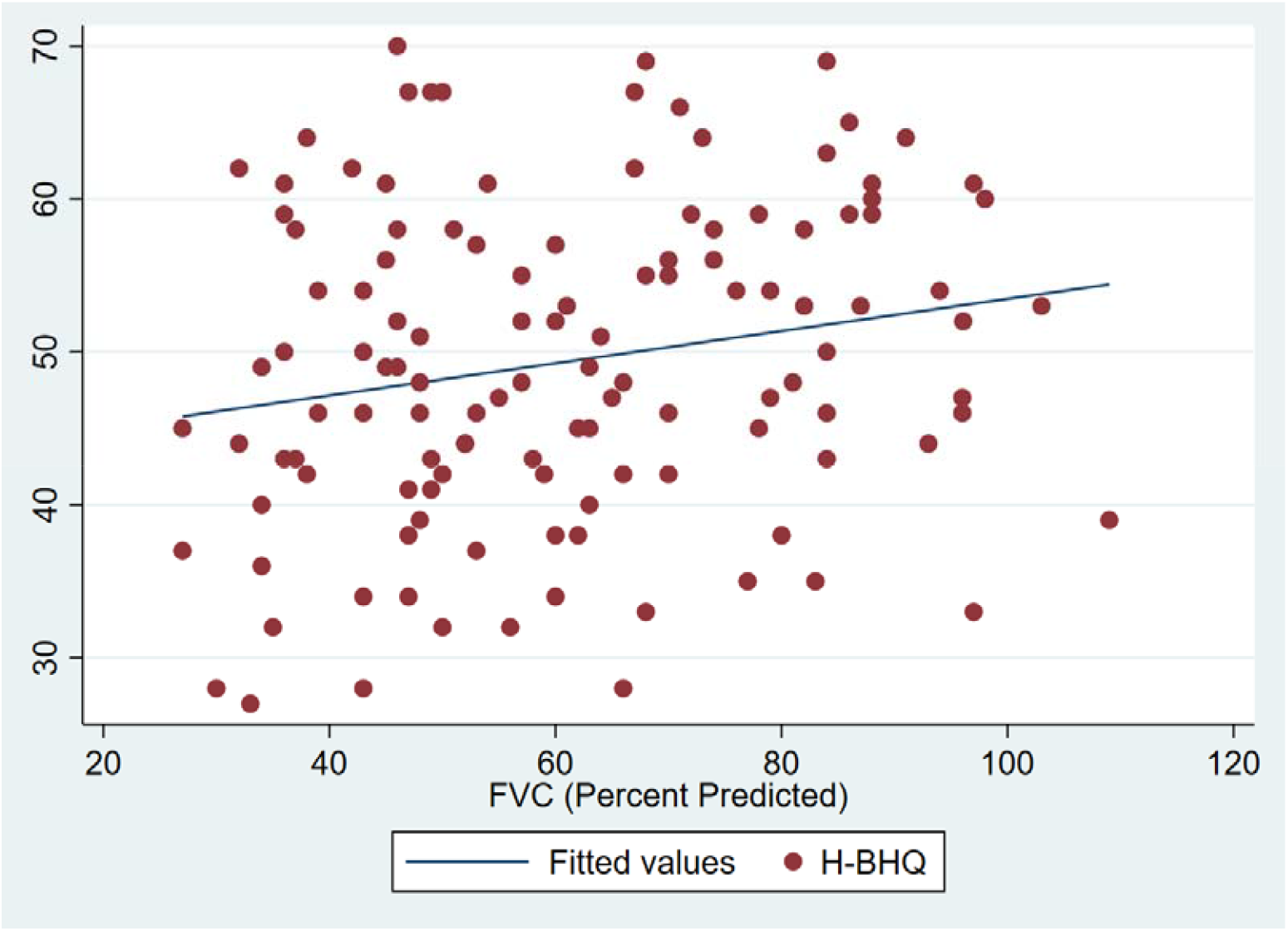
Scatter plot showing the correlation between the H-BHQ scores and the FVC (percent predicted)

## Discussion

In this study, we have attempted to develop and validate a Hindi version of the Bronchiectasis Health Questionnaire (BHQ). Our version showed a moderately strong correlation with difference construct validities (namely the mMRC score and the number of exacerbations in the previous year) and a weak correlation with the number of hospital admissions and the bronchiectasis severity index. The weaker correlation with the number of hospital admissions suggests that the symptom severity and consequently the BHQ score alone might not be a reliable indicator of the risk of exacerbations severe enough to require hospitalization; and that other factors such as duration of disease, persistent colonization with organisms such as *Pseudomonas* or the extent of disease on radiology, likely taken together, might contribute to the risk of admissions as well.

Our study follows other similar studies which have attempted to validate region-specific versions of the BHQ, namely the Turkish and Korean translations^16,17^. The table below shows how our findings compare to these studies:

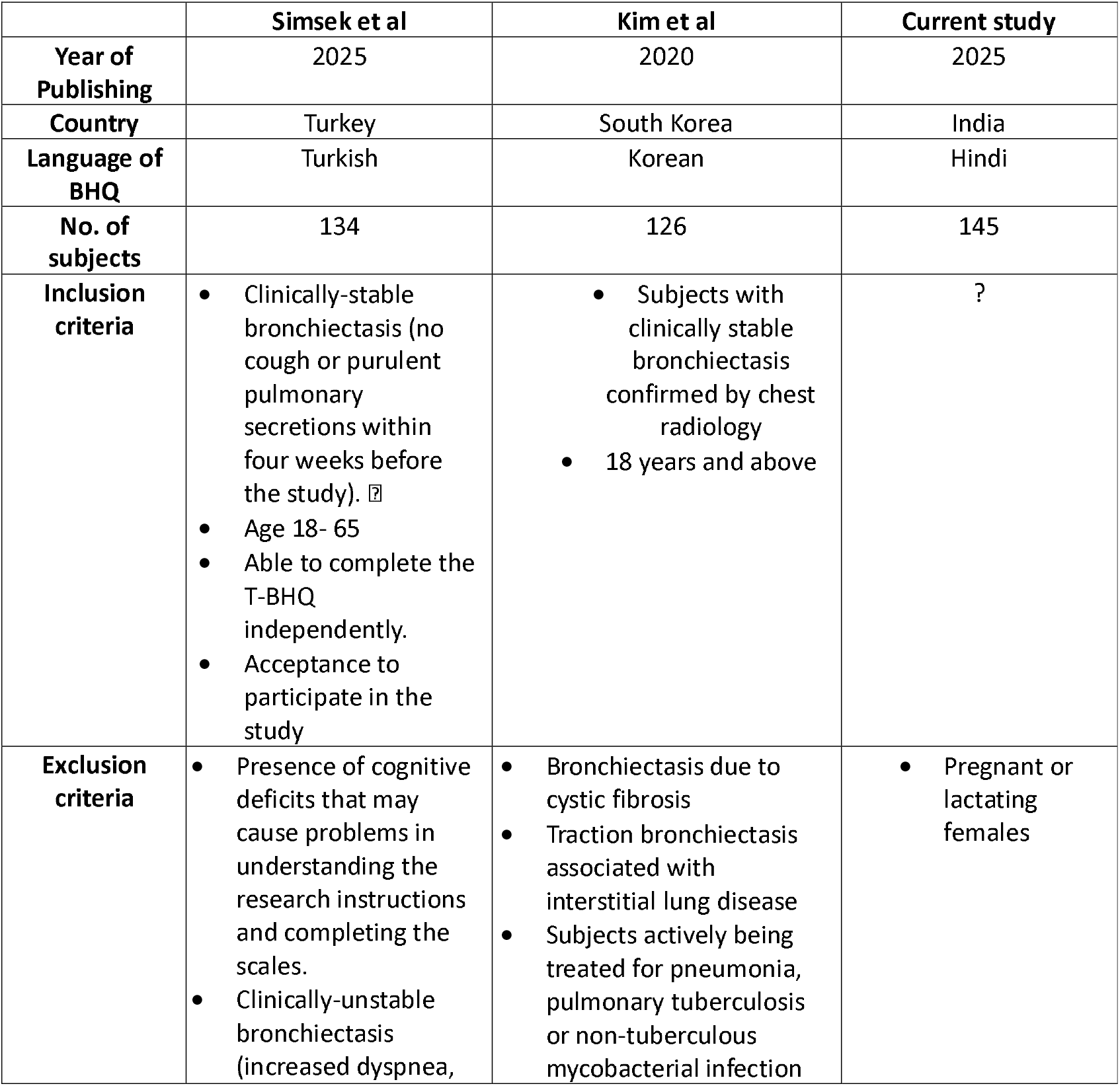
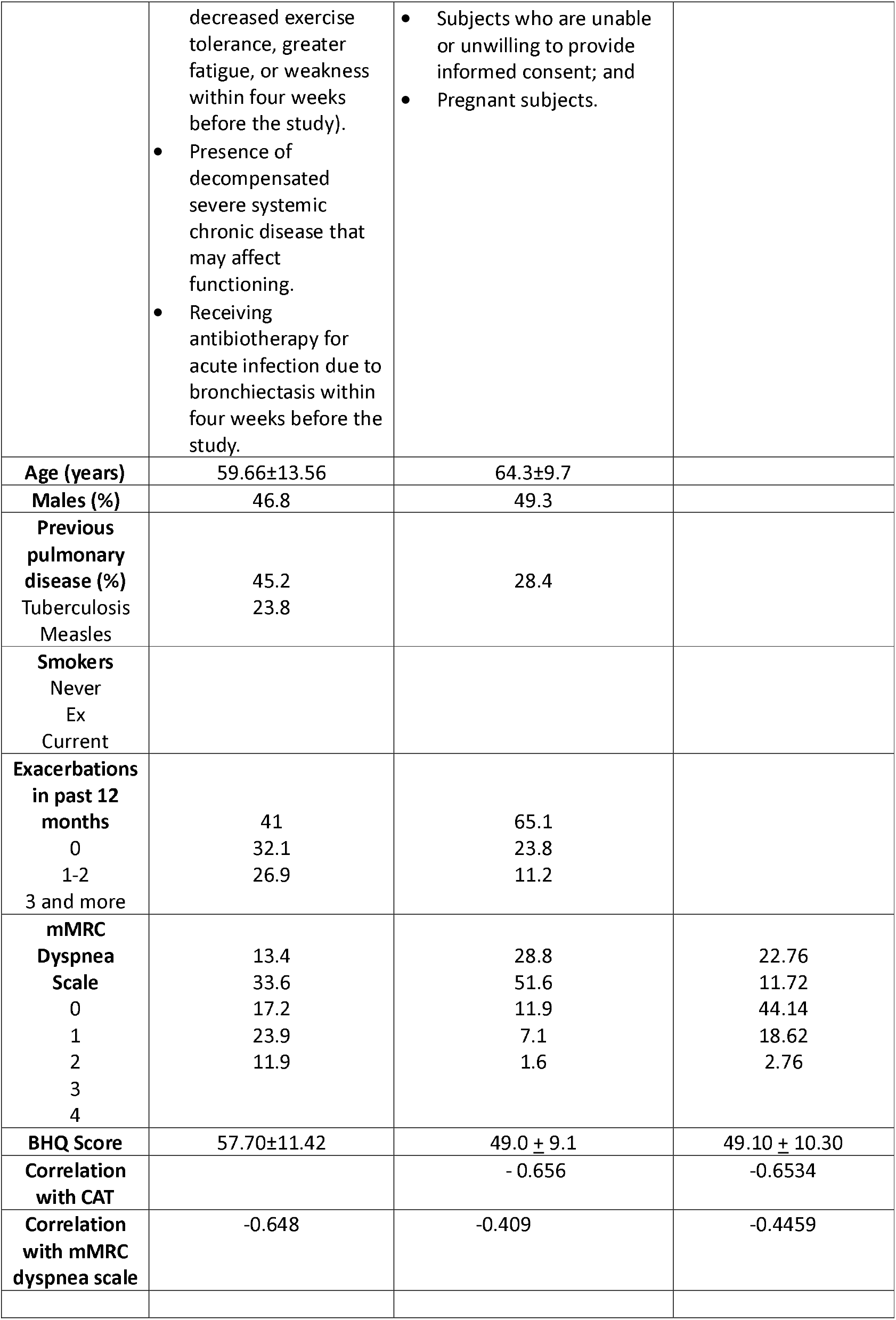

|  | Simsek et al | Kim et al | Current study |
| --- | --- | --- | --- |
| **Year of Publishing** | 2025 | 2020 | 2025 |
| **Country** | Turkey | South Korea | India |
| **Language of BHQ** | Turkish | Korean | Hindi |
| **No. of subjects** | 134 | 126 | 145 |
| **Inclusion criteria** | Clinically-stable bronchiectasis (no cough or purulent pulmonary secretions within four weeks before the study). ☐<br>Age 18- 65<br>Able to complete the T-BHQ independently.<br>Acceptance to participate in the study | Subjects with clinically stable bronchiectasis confirmed by chest radiology<br>18 years and above | ? |
| **Exclusion criteria** | Presence of cognitive deficits that may cause problems in understanding the research instructions and completing the scales.<br>Clinically-unstable bronchiectasis (increased dyspnea, | Bronchiectasis due to cystic fibrosis<br>Traction bronchiectasis associated with interstitial lung disease<br>Subjects actively being treated for pneumonia, pulmonary tuberculosis or non-tuberculous mycobacterial infection | Pregnant or lactating females |
|  | decreased exercise tolerance, greater fatigue, or weakness within four weeks before the study).<br>• Presence of decompensated severe systemic chronic disease that may affect functioning.<br>• Receiving antibiotherapy for acute infection due to bronchiectasis within four weeks before the study. | • Subjects who are unable or unwilling to provide informed consent; and<br>• Pregnant subjects. |  |
| **Age (years)** | 59.66±13.56 | 64.3±9.7 |  |
| **Males (%)** | 46.8 | 49.3 |  |
| **Previous pulmonary disease (%)** |  |  |  |
| Tuberculosis | 45.2 | 28.4 |  |
| Measles | 23.8 |  |  |
| **Smokers** |  |  |  |
| Never |  |  |  |
| Ex |  |  |  |
| Current |  |  |  |
| **Exacerbations in past 12 months** |  |  |  |
| 0 | 41 | 65.1 |  |
| 1-2 | 32.1 | 23.8 |  |
| 3 and more | 26.9 | 11.2 |  |
| **mMRC Dyspnea Scale** |  |  |  |
| 0 | 13.4 | 28.8 | 22.76 |
| 1 | 33.6 | 51.6 | 11.72 |
| 2 | 17.2 | 11.9 | 44.14 |
| 3 | 23.9 | 7.1 | 18.62 |
| 4 | 11.9 | 1.6 | 2.76 |
| **BHQ Score** | 57.70±11.42 | 49.0 ± 9.1 | 49.10 ± 10.30 |
| **Correlation with CAT** |  | - 0.656 | -0.6534 |
| **Correlation with mMRC dyspnea scale** | -0.648 | -0.409 | -0.4459 |

Our study is the second study from the Indian subcontinent to validate the BHQ for chronic respiratory conditions; Jha et al ^18^ in 2025 assessed the psychometric properties of the BHQ in bronchiectasis due to ABPA and reported the meaningful score difference (MSD) in these subjects. The BHQ demonstrated acceptability, strong correlation with Saint George’s Respiratory Questionnaire (r = -0.80), good internal consistency (Cronbach α = 0.81), and test-retest reliability (intraclass correlation coefficient = 0.81). The BHQ scores improved significantly (mean, 57 to 72; P <.001) after treatment. MSD estimates across anchor-based methods ranged from 7.2 to 14.5 points; triangulation yielded an estimate of 10 points. However, this was done using the original English version of the BHQ.

Our study has several strengths. It validated the Hindi version of BHQ in a large Hindi-speaking population presenting to a major tertiary care hospital. Post-tubercular bronchiectasis was the commonest cause of bronchiectasis in this study. India is one of the countries with the highest burden of tuberculosis; consequently, it has one of the largest populations of individuals affected by sequelae of prior tuberculosis, including but not limited to bronchiectasis. However, many subjects seeking healthcare, especially in resource-limited settings, are not able to comprehend English with ease. This makes a Hindi version of commonly used HR-QoL questionnaires such as the BHQ a useful option to have. Further this study had evaluated the correlation of BHQ with most of the important parameters observed in clinical practice. Our study demonstrated moderately strong correlations with the mMRC, CAT scores and exacerbations, which was consistent with results from prior translations of the BHQ. In addition, we also demonstrated significant correlation (though weak) of the H-BHQ with the number of exacerbations requiring hospitalization. Limitations to our study include the absence of cystic fibrosis as an exclusion criterion; treatment guidelines for cystic fibrosis are often distinct from those for bronchiectasis due to other causes.

## Conclusions

The H-BHQ is a questionnaire with significant correlation with mMRC,number of exacerbations in the last year and spirometry parameters like FVC. This questionnaire can be an important adjunct for evaluation of Hind-speaking bronchiectasis subjects.

## Data Availability

All data produced in the present study are available upon reasonable request to the authors

## Abbreviations

aHR: Adjusted Hazard Ratio
BHQ: Bronchiectasis Health Questionnaire
BMI: Body Mass Index
BSI: Bronchiectasis Severity Index
CI: Confidence Interval
CT: Computed Tomography
FEV1: Forced Expiratory Volume in 1 second
FVC: Forced Vital Capacity
H-BHQ: Hindi version of the Bronchiectasis Health Questionnaire
H-CAT: Hindi version of the COPD Assessment Tool
HR-QoL: Health-Related Quality of Life
IgA, IgE, IgG, IgM: Immunoglobulins A, E, G and M
LCQ: Leicester Cough Questionnaire
mMRC: Modified Medical Research Council
PCD: Primary Ciliary Dyskinesia
PICADAR: Primary Ciliary Dyskinesia Rule
PFT: Pulmonary Function Testing
SD: Standard Deviation
SGRQ: St. George’s Respiratory Questionnaire
TB: Tuberculosis
US: United States
USD: United States Dollar

